# Non-English Communication Access in Mental Health and Substance Use Treatment Facilities in the U.S., 2024

**DOI:** 10.1101/2025.11.18.25340414

**Authors:** Alejandro J. Faria, Meagan K. Sullivan, Tyler G. James

**Affiliations:** Unaffiliated investigator, Gainesville, FL; Department of Family Medicine, University of Michigan, Ann Arbor, MI; Department of Psychology, University of Florida, Gainesville, FL

**Keywords:** Limited English Proficiency, Non-English speaking, mental health, Patient Protection and Affordable Care Act, Section 1557, patient-provider communication

## Abstract

Section 1557 of the Patient Protection and Affordable Care Act requires language access services in healthcare organizations receiving federal monies. In 2024, 32.0 percent of publicly funded mental health facilities and 39.9 percent of publicly funded substance use treatment facilities were in non-compliance with Section 1557. We present findings from across the United States using GIS mapping. We also present information on disparities in access to direct counseling in a non-English language.

## Introduction

Non-English-speaking populations, or those with limited English proficiency (LEP), in the United States are frequently at higher risk of deleterious social determinants of health, and face discrimination in society and healthcare.^1–6^ In the U.S., LEP is a primary risk factor for poor mental health, hindering healthcare navigation and communication with providers.^2–5,7–9^ LEP populations face further obstacles to healthcare access including cultural stigma towards healthcare, geographic availability of resources, racial prejudice, insufficient multicultural training for healthcare professionals, and socioeconomic position^2–5,7–9^; these obstacles increase the risk of poorer health status. Therefore, addressing cultural competency and language access in healthcare is necessary to improve access to care for this population.

Linguistically-concordant care and trained language interpreters are paramount to the successful delivery of services for LEP clients.^1,4,6–8^ Language access in healthcare facilities— including mental healthcare and substance use treatment facilities—is a protected right in the United States under Section 1557 of the Patient Protection and Affordable Care Act.^6^ Section 1557 requires health organizations receiving federal funds (known as “covered entities”) to comply with non-discrimination provisions; this includes providing interpreters for non-English-speaking patients. Despite this requirement, a study of mental health and substance use facility compliance with Section 1557 for deaf and hard-of-hearing patients who use American Sign Language found 41% of mental health and 59% of substance use treatment facilities non-compliant.^6^ Barriers to providing spoken-language interpreter services include lack of availability, interpreters’ lack of experience working with certain communities, or distrust by immigrant clients with fears of jeopardized confidentiality leading to being reported to their country of origin.^2,4,5,7,8^ Reliance on family members for language facilitation when interpreters are unavailable or not requested leads to worse mental health outcomes, breaches in privacy, miscommunication, family tension, and maladaptive coping behaviors such as substance use.^1,7^

Regulations requires language accommodations for LEP patients in mental health facilities. The purpose of this study is to extend previous research, focused on American Sign Language and deaf patients, to identify prevalence of non-compliance with Section 1557 for non-English spoken-language accommodations in mental health and substance use treatment facilities in the U.S. Specifically, we assessed covered entities under Section 1557 to (1) describe the prevalence of non-compliance and (2) identify the most common non-English languages available for direct counseling. These findings are intended to identify gaps in service provision and opportunities to improve compliance and, through systemic change, health equity.

## Methods

We used data from the 2024 National Substance Use and Mental Health Services Survey (N-SUMHSS) administered by the Substance Abuse and Mental Health Services Administration (SAMHSA). The N-SUMHSS samples all known substance use and mental health facilities in the U.S. Correctional facilities are excluded from the final dataset. In total, ~29,000 facilities were eligible with over 90% responding. Additional information on the N-SUMHSS methods are described elsewhere.^10^

Facilities were classified as covered entities under Section 1557 based on reported receipt of funds from the federal government (i.e., Medicare; Medicaid; Community block grant; Community mental health service block; Federal grant funds; Federal military insurance; Department of Veterans Affairs; Indian Health Service, Tribal Health Services, and Urban Indian Health Programs). Facilities were asked if they provided treatment services in languages other than English. Facilities providing these services were asked if these services were provided by a bilingual staff member, a professional interpreter (including web-based interpreters), or both bilingual staff and interpreters. Facilities then selected the languages in which they provided direct counseling (i.e., without an interpreter). Measured languages include Spanish, other common languages (e.g., French, Korean), or Indigenous or American Indian/Alaskan Native (AIAN) languages (e.g., Hopi, Navajo). Questions regarding which languages were available were segmented based on types of treatment services provided.

Data management and analyses were conducted in SAS version 9.4 (see **Supplemental Appendix 1**). Analyses were stratified by service type (mental health or substance use). Due to branch logic in the survey, substance use treatment facilities that also offered mental health treatment services – but did not identify as combined facilities at the onset – were not asked any questions about language access for mental health services. Therefore, language access estimates for mental health facilities are missing approximately half of eligible facilities in the U.S. We estimated prevalence of non-compliance by facility type and by state, then mapped these proportions using GIS software available from Datawrapper.de. We also created heatmaps to identify coverage of services in non-English languages compared to the language needs of the population in a specific state using *ggplot* in R.

## Limitations

Most limitations are based on the data source and, therefore, cannot be resolved. First, the N-SUMHSS excludes correctional facilities (e.g., jails and prisons). The evidence of mental health disparities in these facilities is well-established^11^, as is the lack of language access.^12^ The survey branching logic also systematically excluded substance use treatment facilities providing mental health treatment services from questions related to language access in mental health programs. Previous data, focused on American Sign Language, before this survey branching was added, found that over half of covered substance use treatment facilities were in non-compliance. Based on the exclusion of correctional facilities and previous findings^6^, we expect that our estimates of non-compliance are underestimated. In addition, due to idiosyncrasies specific to patients and their contexts, we are unable to say that facilities providing services are *compliant*. For example, compliance may not occur for a non-English speaker when an interpreter is unqualified. Lastly, our mapping program does not include U.S. territories, so these locations are not mapped.

## Results

In total, 14,316 mental health facilities and 16,168 substance use treatment facilities were included in the analysis. **Exhibit 1** provides the prevalence of non-compliance with Section 1557 among mental health and substance use treatment facilities, stratified by covered entity status. There was a clear association between covered entity status and providing services in non-English languages, with more non-covered entities being non-compliant; the statistical effect of these associations was weak, but significant (Cramer’s V = 0.20 and 0.18, respectively). Among covered entities, 32.0% of mental health facilities and 39.9% of substance use treatment facilities were in non-compliance. The distribution of non-compliance across states for both mental health and substance use treatment facilities is in **Exhibit 2a and 2b** and in **Supplemental Appendix 2: Table B-2**.

The most common language service providers for both mental health and substance use treatment facilities were on-call interpreters (57.7 and 57.2%, respectively). The most common non-English languages for *direct counseling* (i.e., without an interpreter) in covered mental health facilities were: Spanish (n = 3,455), Creole (n=260), and French (n=255); for substance use, Spanish (n=3,183), French (n=191), and Creole (n=167). The language distribution for direct counseling by facility type and covered entity status is in **Supplemental Appendix 2: Table B-4**, and visualized by state and top three languages in **Supplemental Figure B-1 and B-2**; 40.4% and 44.2% of states and territories were missing at least one top language for direct counseling in covered mental health and substance use treatment facilities, respectively.

Several gaps in access to direct counseling existed particularly for other American Indian and Alaskan Native languages, Vietnamese, German, and French.

## Discussion

For both mental health and substance use treatment facilities, facilities receiving federal funds (i.e., covered entities) were more likely to provide services in a non-English language than facilities not receiving federal funds. The higher availability of non-English services among covered entities may suggest a degree of effectiveness for federal non-discrimination requirements in protecting patients’ civil rights within healthcare. Opportunity for improvement among covered entities remains evident, as over a quarter of covered mental health facilities and almost two-fifths of covered substance use treatment facilities were identified as non-compliant. These data indicate the establishment of non-discrimination requirements does not guarantee compliance.

Among facilities that provide services in non-English languages, most covered entities provided only on-call interpreters for mental health and substance use treatment. The proportion of facilities providing both bilingual staff and on-call interpreters was slightly higher for mental health facilities than for substance use treatment facilities, across both covered and non-covered entities. Interestingly, significantly more non-covered entities provided *only* bilingual staff across both mental health (28.2%) and substance use treatment (39.9%) compared to covered entities (9.0% and 11.5% for mental health and substance use treatment facilities, respectively). Additional studies identifying why this difference exists would be illuminating for improving policy development, implementation, and enforcement.

Although direct counseling in a patient’s preferred language is considered best practice for providing care for LEP populations,^1,4,6,7^ there were considerable gaps in access to direct counseling. This illustrates a disconnect between the most common languages spoken by the population in the state and the direct counseling services available. Reliance on on-call interpreters alone increases the risk of miscommunication and misunderstanding between providers and LEP patients.^4,7^ Studies investigating facilitators and barriers to providing direct counseling in prevalent non-English languages within mental health and substance use facilities would inform interventions for improving access to appropriate diagnosis and care for LEP populations.

### List of Exhibits

EXHIBIT 1 (table)

Caption: Prevalence of non-compliance among mental health and substance use treatment facilities in the U.S., by covered entity status under the Affordable Care Act Section 1557, 2024. Source/Notes: SOURCE Author’s analysis of N-SUMHSS data.

NOTES. ^a^ Among facilities that indicate providing services in non-English language.

EXHIBIT 2a (figure)

Caption: Prevalence of non-compliance among mental health treatment facilities in the U.S., among facilities federally required by Section 1557 to provide language services, 2024.

Source/Notes: SOURCE Author’s analysis of N-SUMHSS data.

EXHIBIT 2b (figure)

Caption: Prevalence of non-compliance among substance use treatment facilities in the U.S., among facilities federally required by Section 1557 to provide language services, 2024.

Source/Notes: SOURCE Author’s analysis of N-SUMHSS data.

**Exhibit 1.** Prevalence of non-compliance among mental health and substance use treatment facilities in the U.S., by covered entity status under the Affordable Care Act Section 1557, 2024.

|  | Mental health |  | Substance use |  |
| --- | --- | --- | --- | --- |
|  | Not covered entity | Covered entity | Not covered entity | Covered entity |
| Facility does not provide services in non-English language | 70.5% (670) | 32.0% (4281) | 66.6% (1293) | 39.9% (5672) |
| Facility provides services in non-English language | 29.5% (280) | 68.0% (9085) | 33.4% (647) | 60.1% (8556) |
| **Language service provider<sup>a</sup>** |  |  |  |  |
| Bilingual staff | 28.2% (79) | 9.0% (815) | 39.9% (294) | 11.5% (980) |
| On-call interpreters | 37.9% (106) | 57.7% (5235) | 33.1% (204) | 57.2% (4902) |
| Both bilingual staff and on-call interpreters | 33.9% (95) | 33.4% (3028) | 26.9% (195) | 31.1% (2655) |

<sup>a</sup> Among facilities that indicate providing services in non-English language.

**Exhibit 2a.**
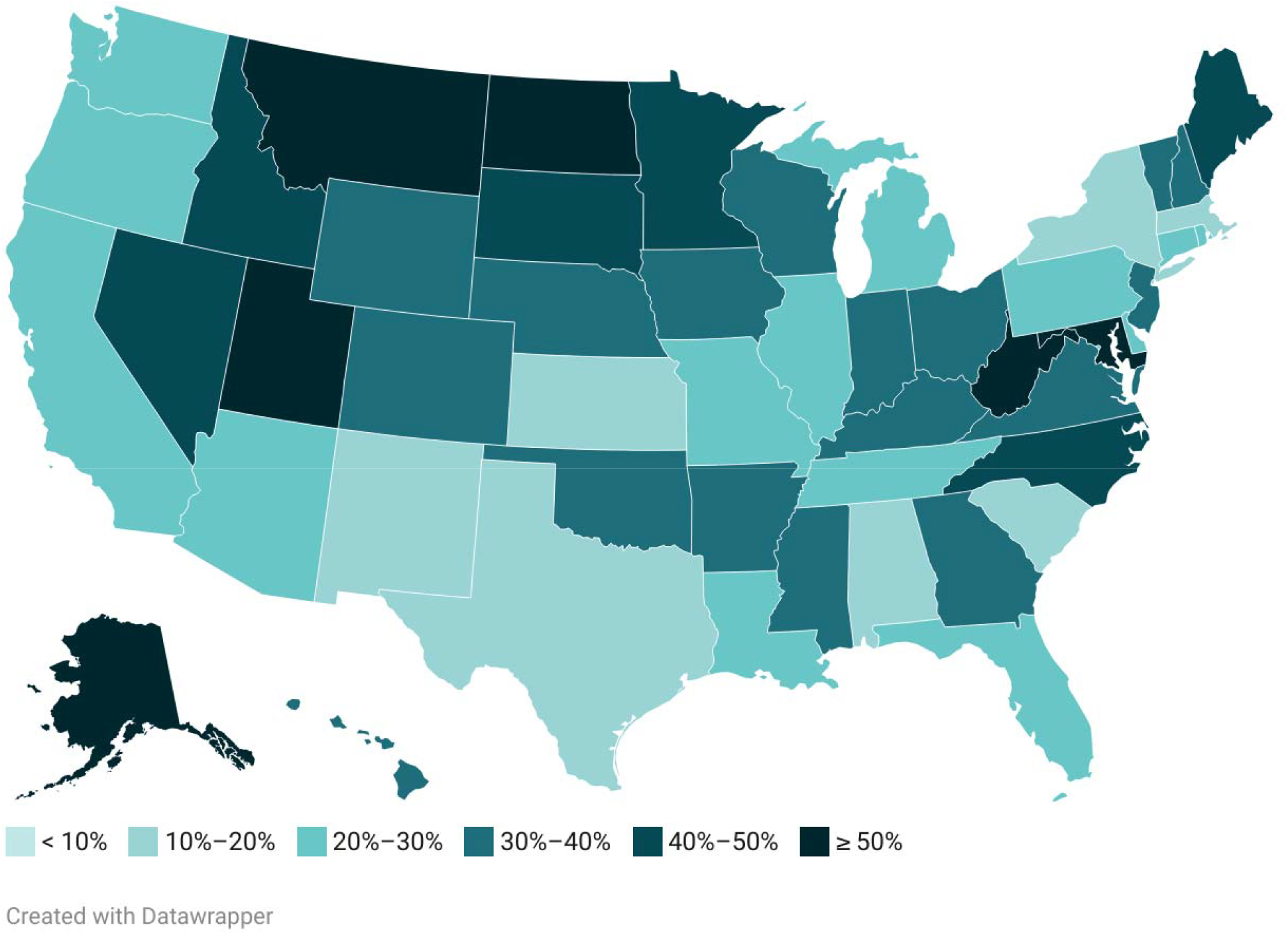
Prevalence of non-compliance among mental health treatment facilities in the U.S., among facilities federally required by Section 1557 to provide language services, 2024.

**Exhibit 2b.**
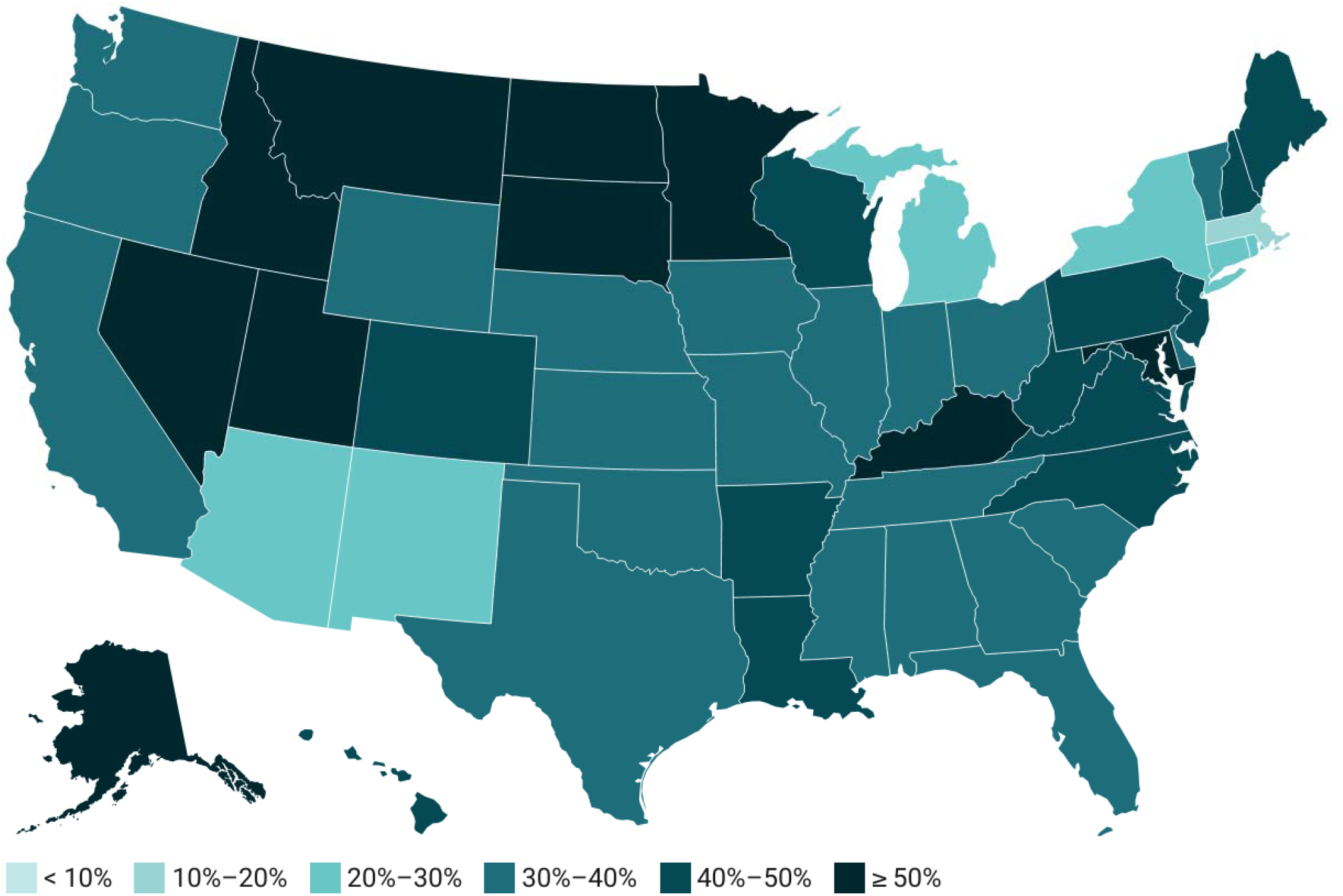
Prevalence of non-compliance among substance u se treatment facilities in the U.S., among facilities federally required by Section 1557 to provide language services, 2024.

## Data Availability

Data are publicly available from the U.S. Substance Abuse and Mental Health Services Administration (SAMHSA).

https://www.samhsa.gov/data/data-we-collect/n-sumhss-national-substance-use-and-mental-health-services-survey/datafiles

