## Supplemental Appendix for "Non-English Communication Access in Mental Health and Substance Use Treatment Facilities in the U.S., 2024"

**Supplemental Appendix 1.** Statistical code for SAS.

```
/* Preparation */

*Library;
LIBNAME AJF "Z:\NSUMHSS Faria";

*Load in clean dataset;
DATA AJFDATAUSE;
	SET AJF.WORKNSUMHSS;
RUN;

/* Facility characteristics */

TITLE1 "FACILITY CHARACTERISTICS - FOCUS AREAS, RECODED";
PROC FREQ DATA = AJFDATAUSE;
TABLE
n_focus2;
RUN; TITLE1;

/* RQ 1 */

TITLE1 "RQ 1. PREVALENCE OF (NON)COMPLIANCE";

TITLE2 "Mental health facilities, column percents";
PROC FREQ DATA = AJFDATAUSE;
TABLE
lang_mh1*coveredent/norow nopercent chisq;
WHERE (MHYES = 1);
RUN; TITLE2;

TITLE2 "Substance use facilities, column percents";
PROC FREQ DATA = AJFDATAUSE;
TABLE
lang_su1*coveredent/norow nopercent chisq;
WHERE (SUBUSEYES = 1);
RUN; TITLE2;

TITLE2 "Supplement: Mental health facilities, column percents, w/ missing";
PROC FREQ DATA = AJFDATAUSE;
TABLE
lang_mh1*coveredent/norow nopercent;
WHERE (MHYES = 1);
RUN; TITLE2;

TITLE2 "Supplement: Substance use facilities, column percents, w/ missing";
PROC FREQ DATA = AJFDATAUSE;
TABLE
lang_su1*coveredent/norow nopercent;
WHERE (SUBUSEYES = 1);
RUN; TITLE2;

TITLE2 "Supplement: Total mental health facilities by state";
PROC FREQ DATA = AJFDATAUSE;
TABLE
locationstate;
WHERE (MHYES = 1) AND (LANG_MH1 ne .);
RUN; TITLE2;

TITLE2 "Supplement: Mental health facilities, row percents, by state";
PROC TABULATE DATA = AJFDATAUSE;
CLASS LOCATIONSTATE coveredent lang_mh1;
TABLE LOCATIONSTATE, coveredent*lang_mh1;
WHERE MHYES = 1;
RUN; TITLE2;

TITLE2 "Supplement: Total substance use facilities by state";
PROC FREQ DATA = AJFDATAUSE;
TABLE
locationstate;
WHERE (SUBUSEYES = 1) AND (lang_su1 ne .);
RUN; TITLE2;

TITLE2 "Supplement: Substance use facilities, row percents, by state";
PROC TABULATE DATA = AJFDATAUSE;
CLASS LOCATIONSTATE coveredent lang_su1;
TABLE LOCATIONSTATE, coveredent*lang_su1;
WHERE SUBUSEYES = 1;
RUN; TITLE2;

TITLE1;

/* RQ 2 */
/* Among Section 1557 covered entities, what percentage of facilities provide
direct counseling (i.e., without an interpreter) in a non-English
language (e.g., Spanish)? */

TITLE1 "RQ 2. PREVALENCE OF DIRECT COUNSELING IN NON-ENG LANGUAGE";

TITLE2 "Type of language service - mental health facilities";
PROC FREQ DATA = AJFDATAUSE;
TABLE
n_LANGPROV_MH*COVEREDENT/norow nopercent;
WHERE (MHYES = 1);
RUN; TITLE2;

TITLE2 "Type of language service - substance use facilities";
PROC FREQ DATA = AJFDATAUSE;
TABLE
n_LANGPROV_SU*COVEREDENT/norow nopercent;
WHERE (SUBUSEYES = 1);
RUN; TITLE2;

TITLE1;

/* RQ 3 */
/* Among Section 1557 covered entities that provide direct counseling
(i.e., without an interpreter) in a non-English language, what
languages are most prevalent? */

TITLE1 "RQ 3. PREVALENCE OF LANGUAGES USED FOR DIRECT COUNSELING";

/* Prepare for analysis by getting number of languages for mental health */
DATA LONG_MH; SET AJFDATAUSE;

IF(MHYES = 1); /*use denominator*/

ARRAY lang {*}
LANG1N_MH LANG2N_MH LANG3N_MH LANG4N_MH LANG5N_MH LANG6N_MH
LANG7N_MH LANG8N_MH LANG9N_MH LANG10N_MH LANG11N_MH LANG12N_MH
LANG13N_MH LANG14N_MH LANG15N_MH LANG16N_MH LANG17N_MH LANG18N_MH
LANG20N_MH LANG21N_MH LANG22N_MH LANG23N_MH LANG24N_MH LANG25N_MH
LANG26N_MH;

DO i = 1 to dim(lang);
LANG_VAR = vname(lang{i}); /*store variable name*/
LANG_VAL = lang{i}; /*store value*/
IF LANG_VAL = 1 THEN OUTPUT; /*only include LANG#N_SU = 1 */
END;

KEEP LOCATIONSTATE LANG_VAR LANG_VAL COVEREDENT;

RUN;

TITLE2 "Prevalence of languages among mental health facilities";
PROC FREQ DATA = LONG_MH;
TABLE
LANG_VAR*COVEREDENT/nocol nopercent;
RUN; TITLE2;

TITLE2 "Languages for direct counseling, mental health, covered entities, by state";
PROC FREQ DATA = LONG_MH;
TABLE
LOCATIONSTATE*LANG_VAR/nocol norow nopercent out=mh_freq_covered;
WHERE COVEREDENT = 1;
RUN; TITLE2;

/* Expore to CSV for mapping */
PROC EXPORT DATA=mh_freq_covered
OUTFILE="Z:\NSUMHSS Faria\mh_lang_state_covered.csv"
DBMS=CSV REPLACE;
RUN;


/* Prepare for analysis by getting number of languages for mental health, by state*/
DATA LONG_SU; SET AJFDATAUSE;

IF(SUBUSEYES = 1); /*use denominator*/

ARRAY lang {*}
LANG1N_SU LANG2N_SU LANG3N_SU LANG4N_SU LANG5N_SU LANG6N_SU
LANG7N_SU LANG8N_SU LANG9N_SU LANG10N_SU LANG11N_SU LANG12N_SU
LANG13N_SU LANG14N_SU LANG15N_SU LANG16N_SU LANG17N_SU LANG18N_SU
LANG20N_SU LANG21N_SU LANG22N_SU LANG23N_SU LANG24N_SU LANG25N_SU
LANG26N_SU;

DO i = 1 to dim(lang);
LANG_VAR = vname(lang{i}); /*store variable name*/
LANG_VAL = lang{i}; /*store value*/
IF LANG_VAL = 1 THEN OUTPUT; /*only include LANG#N_SU = 1 */
END;

KEEP LOCATIONSTATE LANG_VAR LANG_VAL COVEREDENT;
RUN;

TITLE2 "Prevalence of languages among substance use facilities";
PROC FREQ DATA = LONG_SU;
TABLE
LANG_VAR*COVEREDENT/nocol nopercent;
RUN; TITLE2;

TITLE2 "Languages for direct counseling, substance use, covered entities, by state";
PROC FREQ DATA = LONG_SU;
TABLE LOCATIONSTATE*LANG_VAR/nocol nopercent out=su_freq_covered;
WHERE COVEREDENT = 1;
RUN; TITLE2;

/* Expore to CSV for mapping */
PROC EXPORT DATA=su_freq_covered
OUTFILE="Z:\NSUMHSS Faria\su_lang_state_covered.csv"
DBMS=CSV REPLACE;
RUN; TITLE1;
```

**Supplemental Appendix B. Additional Statistical Results**

**Table B-1.** Prevalence of non-compliance among mental health and substance use treatment facilities in the U.S., by covered entity status under the Affordable Care Act Section 1557, 2024.

|  | **Mental health** | | **Substance use** | |
| --- | --- | --- | --- | --- |
|  | **Not covered entity** | **Covered entity** | **Not covered entity** | **Covered entity** |
| **Missing** | 51.6% (1014) | 25.3% (4519) | 1.5% (30) | 2.5% (361) |
| **No services in non-English language** | 34.1% (670) | 23.9% (4281) | 65.6% (1293) | 38.9% (5672) |
| **Services in non-English language** | 14.3% (280) | 50.8% (9085) | 32.8% (647) | 58.6% (8556) |

*Note*. A substantial number of mental health facilities were missing language service data due to branching logic on the N-SUMHSS. Specifically, substance use treatment facilities that also provide mental health services – that did not self-identify as combined (substance use + mental health) facilities at the beginning of the survey – were not shown the mental health service questions.

**Supplemental Table B-2.** Prevalence of compliant **mental health** facilities by covered entity status by state.

| State | Non-covered entity | | Covered entity | | Total | | |
| --- | --- | --- | --- | --- | --- | --- | --- |
|  | **Not compliant** | **Compliant** | **Not compliant** | **Compliant** | **n** | **% non-compliant, all facilities** | **% non-complaint, covered entities** |
| Alabama | 3 | 0 | 22 | 103 | 128 | 19.5% | 17.6% |
| Alaska | 1 | 0 | 54 | 46 | 101 | 54.5% | 54.0% |
| Arizona | 21 | 4 | 138 | 363 | 526 | 30.2% | 27.5% |
| Arkansas | 6 | 0 | 78 | 131 | 215 | 39.1% | 37.3% |
| California | 185 | 83 | 273 | 685 | 1226 | 37.4% | 28.5% |
| Colorado | 19 | 5 | 65 | 136 | 225 | 37.3% | 32.3% |
| Connecticut | 8 | 2 | 55 | 179 | 244 | 25.8% | 23.5% |
| Delaware | 6 | 0 | 9 | 35 | 50 | 30.0% | 20.5% |
| District of Columbia | 0 | 0 | 5 | 14 | 19 | 26.3% | 26.3% |
| Florida | 53 | 29 | 141 | 437 | 660 | 29.4% | 24.4% |
| Georgia | 20 | 4 | 69 | 157 | 250 | 35.6% | 30.5% |
| Hawaii | 0 | 4 | 12 | 23 | 39 | 30.8% | 34.3% |
| Idaho | 1 | 1 | 59 | 61 | 122 | 49.2% | 49.2% |
| Illinois | 35 | 17 | 112 | 347 | 511 | 28.8% | 24.4% |
| Indiana | 7 | 4 | 124 | 277 | 412 | 31.8% | 30.9% |
| Iowa | 1 | 2 | 61 | 105 | 169 | 36.7% | 36.7% |
| Kansas | 1 | 1 | 22 | 110 | 134 | 17.2% | 16.7% |
| Kentucky | 7 | 3 | 142 | 225 | 377 | 39.5% | 38.7% |
| Louisiana | 6 | 0 | 50 | 121 | 177 | 31.6% | 29.2% |
| Maine | 1 | 0 | 74 | 97 | 172 | 43.6% | 43.3% |
| Maryland | 10 | 3 | 246 | 183 | 442 | 57.9% | 57.3% |
| Massachusetts | 13 | 12 | 52 | 254 | 331 | 19.6% | 17.0% |
| Michigan | 6 | 1 | 101 | 319 | 427 | 25.1% | 24.0% |
| Minnesota | 5 | 4 | 179 | 221 | 409 | 45.0% | 44.8% |
| Mississippi | 7 | 1 | 48 | 94 | 150 | 36.7% | 33.8% |
| Missouri | 5 | 9 | 73 | 193 | 280 | 27.9% | 27.4% |
| Montana | 2 | 0 | 64 | 32 | 98 | 67.3% | 66.7% |
| Nebraska | 4 | 1 | 54 | 86 | 145 | 40.0% | 38.6% |
| Nevada | 3 | 1 | 50 | 72 | 126 | 42.1% | 41.0% |
| New Hampshire | 3 | 0 | 26 | 45 | 74 | 39.2% | 36.6% |
| New Jersey | 23 | 6 | 132 | 199 | 360 | 43.1% | 39.9% |
| New Mexico | 1 | 0 | 24 | 97 | 122 | 20.5% | 19.8% |
| New York | 6 | 11 | 150 | 637 | 804 | 19.4% | 19.1% |
| North Carolina | 18 | 3 | 182 | 203 | 406 | 49.3% | 47.3% |
| North Dakota | 0 | 3 | 22 | 20 | 45 | 48.9% | 52.4% |
| Ohio | 13 | 2 | 261 | 482 | 758 | 36.1% | 35.1% |
| Oklahoma | 1 | 0 | 53 | 115 | 169 | 32.0% | 31.5% |
| Oregon | 0 | 1 | 58 | 155 | 214 | 27.1% | 27.2% |
| Pennsylvania | 33 | 10 | 145 | 340 | 528 | 33.7% | 29.9% |
| Puerto Rico | 1 | 9 | 1 | 51 | 62 | 3.2% | 1.9% |
| Rhode Island | 0 | 0 | 11 | 44 | 55 | 20.0% | 20.0% |
| South Carolina | 1 | 0 | 13 | 81 | 95 | 14.7% | 13.8% |
| South Dakota | 0 | 0 | 20 | 27 | 47 | 42.6% | 42.6% |
| Tennessee | 18 | 6 | 84 | 206 | 314 | 32.5% | 29.0% |
| Texas | 23 | 19 | 76 | 332 | 450 | 22.0% | 18.6% |
| Utah | 43 | 12 | 157 | 148 | 360 | 55.6% | 51.5% |
| Vermont | 4 | 0 | 21 | 41 | 66 | 37.9% | 33.9% |
| Virginia | 18 | 4 | 113 | 185 | 320 | 40.9% | 37.9% |
| Washington | 21 | 1 | 98 | 267 | 387 | 30.7% | 26.8% |
| West Virginia | 3 | 0 | 74 | 62 | 139 | 55.4% | 54.4% |
| Wisconsin | 4 | 2 | 111 | 202 | 319 | 36.1% | 35.5% |
| Wyoming | 0 | 0 | 16 | 35 | 51 | 31.4% | 31.4% |
| Other jurisdiction | 0 | 0 | 1 | 5 | 6 | 16.7% | 16.7% |
| Total | 670 | 280 | 4281 | 9085 | 14316 | 34.6% | 32.0% |

*Note.* Excludes cases missing on language status.

**Supplemental Table B-3.** Prevalence of compliant **substance use** facilities by covered entity status by state.

| State | Non-covered entity | | Covered entity | | Total | | |
| --- | --- | --- | --- | --- | --- | --- | --- |
|  | **Not compliant** | **Compliant** | **Not compliant** | **Compliant** | **n** | **% non-compliant, all facilities** | **% non-complaint, covered entities** |
| Alabama | 25 | 9 | 45 | 74 | 153 | 45.8% | 37.8% |
| Alaska | 0 | 2 | 57 | 31 | 90 | 63.3% | 64.8% |
| Arizona | 25 | 13 | 135 | 371 | 544 | 29.4% | 26.7% |
| Arkansas | 11 | 4 | 57 | 84 | 156 | 43.6% | 40.4% |
| California | 230 | 144 | 449 | 684 | 1507 | 45.1% | 39.6% |
| Colorado | 49 | 15 | 111 | 156 | 331 | 48.3% | 41.6% |
| Connecticut | 9 | 1 | 44 | 138 | 192 | 27.6% | 24.2% |
| Delaware | 9 | 8 | 15 | 31 | 63 | 38.1% | 32.6% |
| District of Columbia | 0 | 0 | 12 | 17 | 29 | 41.4% | 41.4% |
| Florida | 100 | 62 | 187 | 356 | 705 | 40.7% | 34.4% |
| Georgia | 76 | 15 | 98 | 163 | 352 | 49.4% | 37.5% |
| Hawaii | 25 | 35 | 23 | 24 | 107 | 44.9% | 48.9% |
| Idaho | 1 | 0 | 82 | 27 | 110 | 75.5% | 75.2% |
| Illinois | 75 | 58 | 185 | 337 | 655 | 39.7% | 35.4% |
| Indiana | 25 | 12 | 157 | 306 | 500 | 36.4% | 33.9% |
| Iowa | 3 | 0 | 76 | 121 | 200 | 39.5% | 38.6% |
| Kansas | 12 | 4 | 56 | 101 | 173 | 39.3% | 35.7% |
| Kentucky | 29 | 18 | 279 | 271 | 597 | 51.6% | 50.7% |
| Louisiana | 23 | 2 | 85 | 95 | 205 | 52.7% | 47.2% |
| Maine | 5 | 0 | 63 | 82 | 150 | 45.3% | 43.4% |
| Maryland | 17 | 4 | 364 | 187 | 572 | 66.6% | 66.1% |
| Massachusetts | 13 | 9 | 72 | 340 | 434 | 19.6% | 17.5% |
| Michigan | 19 | 2 | 126 | 307 | 454 | 31.9% | 29.1% |
| Minnesota | 9 | 3 | 250 | 127 | 389 | 66.6% | 66.3% |
| Mississippi | 13 | 6 | 33 | 66 | 118 | 39.0% | 33.3% |
| Missouri | 27 | 15 | 95 | 157 | 294 | 41.5% | 37.7% |
| Montana | 1 | 0 | 72 | 29 | 102 | 71.6% | 71.3% |
| Nebraska | 5 | 0 | 53 | 80 | 138 | 42.0% | 39.8% |
| Nevada | 7 | 6 | 63 | 59 | 135 | 51.9% | 51.6% |
| New Hampshire | 5 | 0 | 40 | 49 | 94 | 47.9% | 44.9% |
| New Jersey | 65 | 19 | 146 | 184 | 414 | 51.0% | 44.2% |
| New Mexico | 2 | 6 | 29 | 112 | 149 | 20.8% | 20.6% |
| New York | 27 | 14 | 187 | 619 | 847 | 25.3% | 23.2% |
| North Carolina | 87 | 16 | 212 | 218 | 533 | 56.1% | 49.3% |
| North Dakota | 5 | 7 | 43 | 14 | 69 | 69.6% | 75.4% |
| Ohio | 15 | 5 | 287 | 436 | 743 | 40.6% | 39.7% |
| Oklahoma | 4 | 4 | 55 | 121 | 184 | 32.1% | 31.3% |
| Oregon | 1 | 4 | 78 | 165 | 248 | 31.9% | 32.1% |
| Pennsylvania | 52 | 15 | 254 | 268 | 589 | 52.0% | 48.7% |
| Puerto Rico | 0 | 38 | 1 | 32 | 71 | 1.4% | 3.0% |
| Rhode Island | 0 | 1 | 13 | 47 | 61 | 21.3% | 21.7% |
| South Carolina | 9 | 5 | 27 | 63 | 104 | 34.6% | 30.0% |
| South Dakota | 1 | 2 | 30 | 20 | 53 | 58.5% | 60.0% |
| Tennessee | 39 | 16 | 116 | 214 | 385 | 40.3% | 35.2% |
| Texas | 57 | 23 | 158 | 328 | 566 | 38.0% | 32.5% |
| Utah | 22 | 9 | 140 | 113 | 284 | 57.0% | 55.3% |
| Vermont | 2 | 0 | 21 | 33 | 56 | 41.1% | 38.9% |
| Virginia | 10 | 6 | 162 | 206 | 384 | 44.8% | 44.0% |
| Washington | 24 | 9 | 115 | 226 | 374 | 37.2% | 33.7% |
| West Virginia | 10 | 0 | 70 | 79 | 159 | 50.3% | 47.0% |
| Wisconsin | 11 | 0 | 122 | 149 | 282 | 47.2% | 45.0% |
| Wyoming | 0 | 1 | 21 | 34 | 56 | 37.5% | 38.2% |
| Other jurisdiction | 2 | 0 | 1 | 5 | 8 | 37.5% | 16.7% |
| Total | 1293 | 647 | 5672 | 8556 | 16168 | 43.1% | 39.9% |

**Supplemental Table B-4.** Prevalence of languages used in **direct counseling** in a non-English language.

| Language | Mental health | | | Substance use | | |
| --- | --- | --- | --- | --- | --- | --- |
|  | **Not covered entities** | **Covered entities** | **Total facilities providing language** | **Not covered entities** | **Covered entities** | **Total facilities providing language** |
| American Indian or Alaskan Native languages |  |  |  |  |  |  |
| Hopi [1] | 0 (0%) | 47 (1%) | 47 | 0 (0%) | 34 (1%) | 34 |
| Lakota [2] | 0 (0%) | 41 (1%) | 41 | 0 (0%) | 24 (1%) | 24 |
| Navajo [3] | 0 (0%) | 62 (1%) | 62 | 2 (0.03%) | 64 (0.96%) | 66 |
| Ojibwa [21] | 0 (0%) | 39 (1%) | 39 | 0 (0%) | 24 (1%) | 24 |
| Yupik [4] | 0 (0%) | 38 (1%) | 38 | 0 (0%) | 24 (1%) | 24 |
| Other American Indian/Alaskan Native language [5] | 4 (0.11%) | 30 (0.88%) | 34 | 1 (0.03%) | 29 (0.96%) | 30 |
| Arabic [6] | 2 (0.01%) | 184 (0.98%) | 186 | 3 (0.02%) | 116 (0.97%) | 119 |
| Chinese [7] | 10 (0.04%) | 212 (0.95%) | 222 | 7 (0.06%) | 109 (0.93%) | 116 |
| Creole [8] | 8 (0.02%) | 260 (0.97%) | 268 | 13 (0.07%) | 167 (0.92%) | 180 |
| Farsi [24] | 4 (0.02%) | 184 (0.97%) | 188 | 8 (0.07%) | 96 (0.92%) | 104 |
| French [9] | 6 (0.02%) | 255 (0.97%) | 261 | 18 (0.08%) | 191 (0.91%) | 209 |
| German [10] | 2 (0.01%) | 113 (0.98%) | 115 | 5 (0.06%) | 72 (0.93%) | 77 |
| Greek [22] | 0 (0%) | 72 (1%) | 72 | 6 (0.15%) | 34 (0.85%) | 40 |
| Hebrew [25] | 0 (0%) | 113 (1%) | 113 | 5 (0.06%) | 71 (0.93%) | 76 |
| Hindi [26] | 3 (0.01%) | 211 (0.98%) | 214 | 15 (0.12%) | 108 (0.87%) | 123 |
| Hmong [11] | 1 (0.01%) | 98 (0.98%) | 99 | 2 (0.03%) | 51 (0.96%) | 53 |
| Italian [19] | 0 (0%) | 112 (1%) | 112 | 3 (0.03%) | 81 (0.96%) | 84 |
| Japanese [23] | 0 (0%) | 105 (1%) | 105 | 3 (0.05%) | 50 (0.94%) | 53 |
| Korean [12] | 9 (0.05%) | 143 (0.94%) | 152 | 3 (0.03%) | 73 (0.96%) | 76 |
| Polish [13] | 5 (0.04%) | 108 (0.95%) | 113 | 13 (0.12%) | 92 (0.87%) | 105 |
| Portuguese [14] | 3 (0.01%) | 215 (0.98%) | 218 | 10 (0.07%) | 122 (0.92%) | 132 |
| Russian [15] | 9 (0.03%) | 221 (0.96%) | 230 | 17 (0.09%) | 159 (0.90%) | 176 |
| Spanish [16] | 158 (0.04%) | 3455 (0.95%) | 3613 | 382 (0.10%) | 3183 (0.89%) | 3565 |
| Tagalog [20] | 5 (0.02%) | 190 (0.97%) | 195 | 4 (0.03%) | 102 (0.96%) | 106 |
| Vietnamese [17] | 2 (0.01%) | 148 (0.98%) | 150 | 1 (0.01%) | 75 (0.98%) | 76 |
| Other language [18] | 14 (0.03%) | 435 (0.96%) | 449 | 83 (0.13%) | 543 (0.86%) | 626 |

Row percentages within facility type.

**Supplemental Figure B-1.** Top three non-English languages for **direct counseling in mental health treatment facilities** that are covered entities under Section 1557 of the Affordable Care Act, 2024.

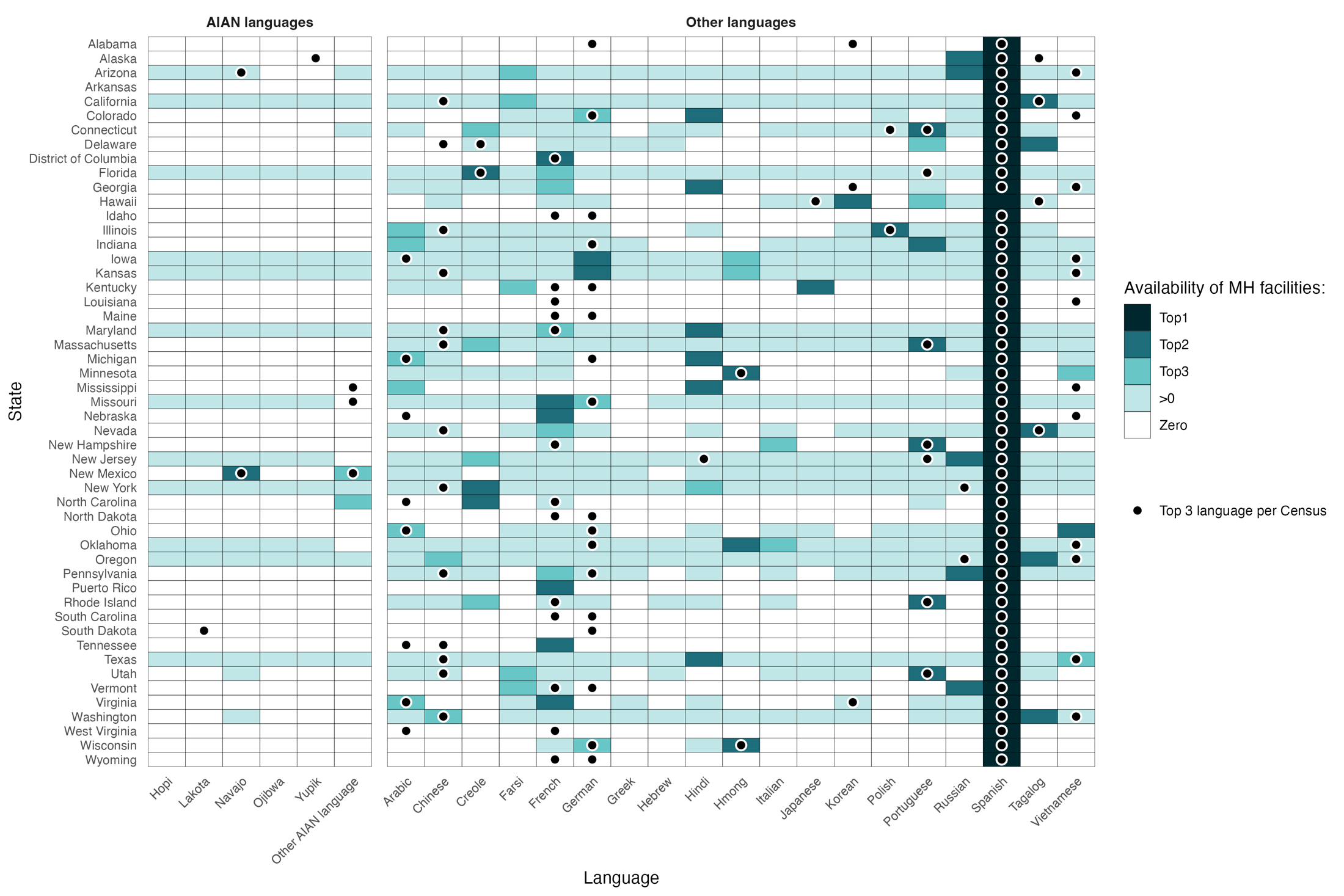

*Notes.* This figure shows the top three languages for direct counseling in mental health treatment facilities by state, with indicators on the top three languages used in the state based on the U.S. Census Bureau. For example, in Colorado, the top three languages used in the state are Spanish, German, and Vietnamese. The most common non-English language offered in mental health facilities for direct counseling was Spanish, followed by Hindi and German; in addition, at least one facility offered direct counseling in Farsi, French, Polish, and Russian. This indicates that there were no publicly funded mental health facilities in Colorado offering direct counseling in Vietnamese, indicating that there was a service gap.

*Data notes.* Top languages are from the per U.S. Census Bureau’s June 2025 tables of *Detailed Languages Spoken at Home and Ability to Speak English for the Population 5 Years and Over: 2017-2021*; this is the most recently available data. There was a lack of data collection for non-English and non-Spanish languages for Puerto Rico. Indiana’s Top 2 and 3 languages are German and Pennsylvania German, combined to ‘German.’ Washington D.C.’s Top 3 language is Amharic. Arkansas’s Top 2 and 3 languages are Marshallese and Lao.

**Supplemental Figure B-2.** Top three non-English languages for **direct counseling** in substance use treatment facilities that are covered entities under Section 1557 of the Affordable Care Act, 2024.

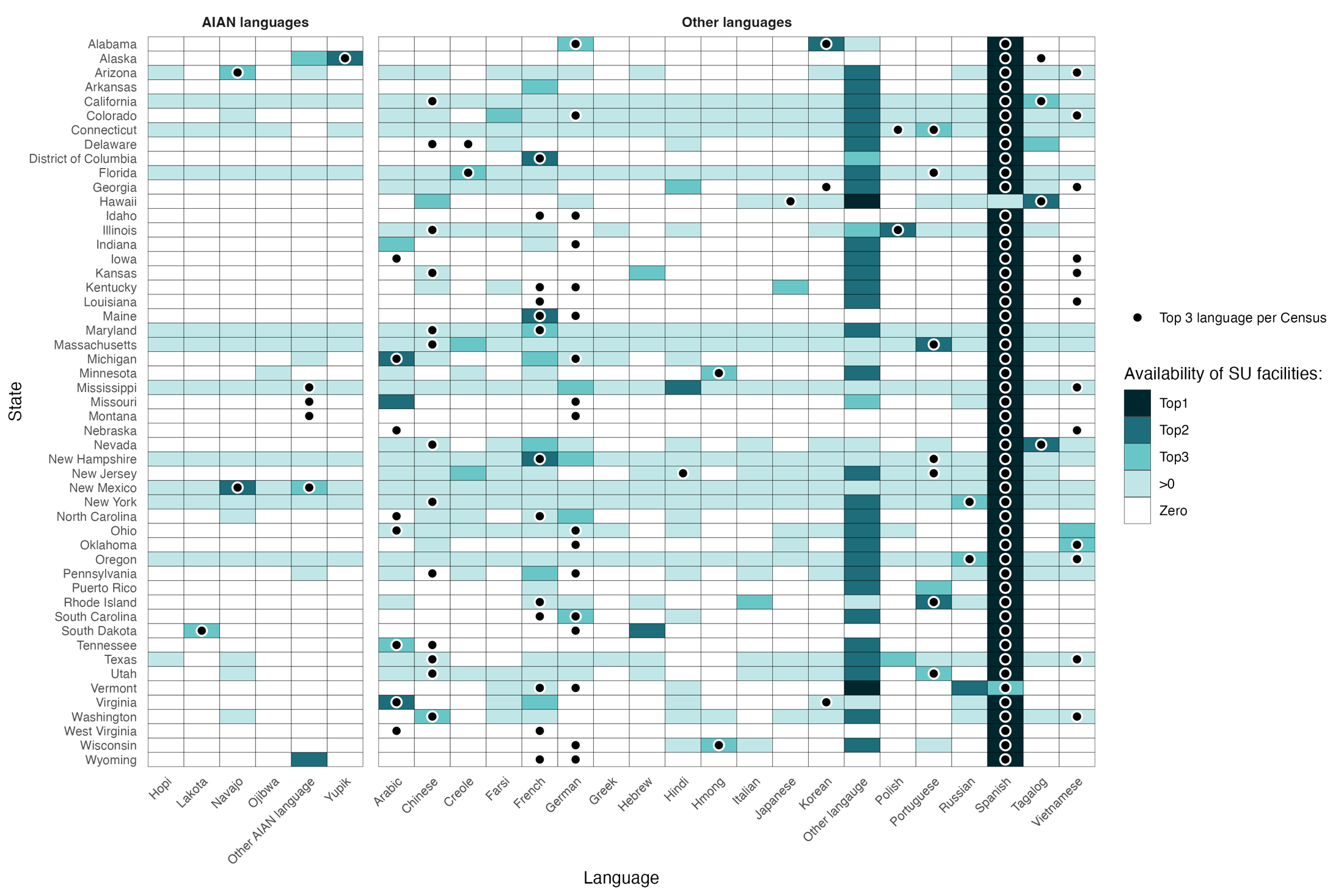

*Notes.* This figure shows the top three languages for direct counseling in substance use treatment facilities by state, with indicators on the top three languages used in the state based on the U.S. Census Bureau. For example, in Alaska, the top three languages used in the state are Spanish, Yupik, and Tagalog. The most common non-English language offered in substance use treatment facilities for direct counseling was Spanish, followed by Yupik and Other AI/AN languages; in addition, no covered entities facility offered direct counseling in any other language surveyed in the N-SUMHSS. This indicates that there were no publicly funded substance use treatment facilities in Alaska offering direct treatment in Tagalog, indicating that there was a service gap.

*Data notes*. Top languages are from the U.S. Census Bureau’s June 2025 tables of *Detailed Languages Spoken at Home and Ability to Speak English for the Population 5 Years and Over: 2017-2021*; this is the most recently available data. There was a lack of data collection for non-English and non-Spanish languages for Puerto Rico. Indiana’s Top 2 and 3 languages are German and Pennsylvania German, combined to ‘German.’ Washington D.C.’s Top 3 language is Amharic. Arkansas’s Top 2 and 3 languages are Marshallese and Lao.
